# Comparison of the Ecological Footprints of administering Salbutamol by Metered-Dose Inhaler and by Nebulization in Emergency Treatment of Acute Asthma

**DOI:** 10.1101/2024.09.22.24314114

**Authors:** Simon Berthelot, Jean-François Ménard, Guillaume Bélanger-Chabot, Gabriela Arias Garcia, Diego Mantovani, Chantale Simard, Jason R. Guertin, Tania Marx, Ariane Bluteau

## Abstract

**Background:** The scientific evidence indicates little or no difference in the effectiveness or cost of using of metered-dose inhalers (MDIs) versus nebulization to treat acute asthma in the emergency department (ED). However, the use of MDIs raises questions of environmental impact. The objective of this study was to compare the carbon footprint of salbutamol administered by MDI versus nebulization.

**Methods:** Applying a life cycle assessment methodology, we quantified the resources extracted and pollutants emitted by each therapeutic option, from the factory production of medication and equipment to disposal by incineration. Each piece of inventory data was then translated into CO_2_-equivalent emissions (CO_2_eq) using the IPCC2021/GWP100 method. Results were estimated for the administration of 1 and 3 treatments of 800 µg of salbutamol by MDI and 5 mg by nebulization (standard doses for adults and children ≥ 24 kg) and compared to the use of a subcompact car.

**Results:** One and three ED-administered treatments with salbutamol emit respectively 1.9 and 4.0 kg of CO_2_eq via MDI versus 0.9 and 1.0 kg via nebulization, which corresponds to 5.5 km and 11.6 km and to 2.7 km and 2.8 km traveled in a subcompact car. Each series of 8 inhalations from an MDI releases 1.1 kg of CO_2_eq due to emission of the hydrofluoroalkane propellent.

**Interpretation:** Considering the absence or minimal difference in clinical effectiveness, this study suggests that nebulization may be a more eco-efficient administration route than MDIs in the emergency treatment of asthma.

**Trail registration:** N/A

**TAKE-HOME POINTS:** *Study question:* What is the ecological footprint of metered-dose inhalers compared to nebulization for administering salbutamol when treating a patient with acute asthma in the emergency department?

*Results:* Nebulization was found to have half the carbon footprint of 1 MDI administration and one quarter of 3 MDI administrations.

*Interpretation:* Implementing low-emission treatment protocols for acute asthma should be one of many avenues to explore to achieve net-zero greenhouse gas emissions in healthcare services.

## INTRODUCTION

Healthcare services are responsible for 4.6% of greenhouse gas emissions worldwide ^1^. The USA and Canada rank first and third, respectively, in per capita healthcare-related emissions of CO_2_ equivalents (CO_2_eq) ^1^. Increasing emissions of greenhouse gases contribute to global warming, of which the negative impact on human health includes an increase in heat-related illnesses and respiratory problems as well as wider spread of infectious diseases ^2^.

The use of metered-dose inhalers (MDIs) to administer bronchodilators (e.g., salbutamol) is considered a significant source of health-service-associated greenhouse gas emissions ^3^. It has been estimated that in the United Kingdom, up to 13% of such emissions result from the use of MDIs to treat asthma or chronic obstructive pulmonary disease (COPD). The main greenhouse gas emitted from these devices is the hydrofluoroalkane (HFA) propellent. Some HFAs have atmospheric warming potentials up to 3000 times that of CO_2_ ^4,5^. Consequently, replacing MDIs with HFA-free administration methods is regarded as desirable whenever possible. However, MDI use has become the standard of care in many emergency departments ^6,7^. Administration of a bronchodilator in the form of fine droplets is the main therapeutic alternative in the hospital setting. This mode, known as nebulization, is said to have equivalent clinical effectiveness ^8^. To our knowledge, its environmental impact has never been evaluated and compared to that of MDIs ^5^.

The objective of this study was to compare the environmental footprints of MDI and nebulization when used to administer salbutamol to adults in emergency departments.

## METHODS

### Design and Setting

This is an observational study comparing MDIs and nebulization as methods of administering salbutamol to asthmatic patients in emergency departments, using a life cycle assessment methodology to calculate the “cradle to grave” environmental footprints (Figure 1). The study was conducted during fiscal year 2022-23 at the CHU de Québec-Université Laval (hereafter the CHU), an academic institution that manages five EDs located throughout Québec (Canada) and receiving more than 240,000 visits annually.

**Figure 1.**
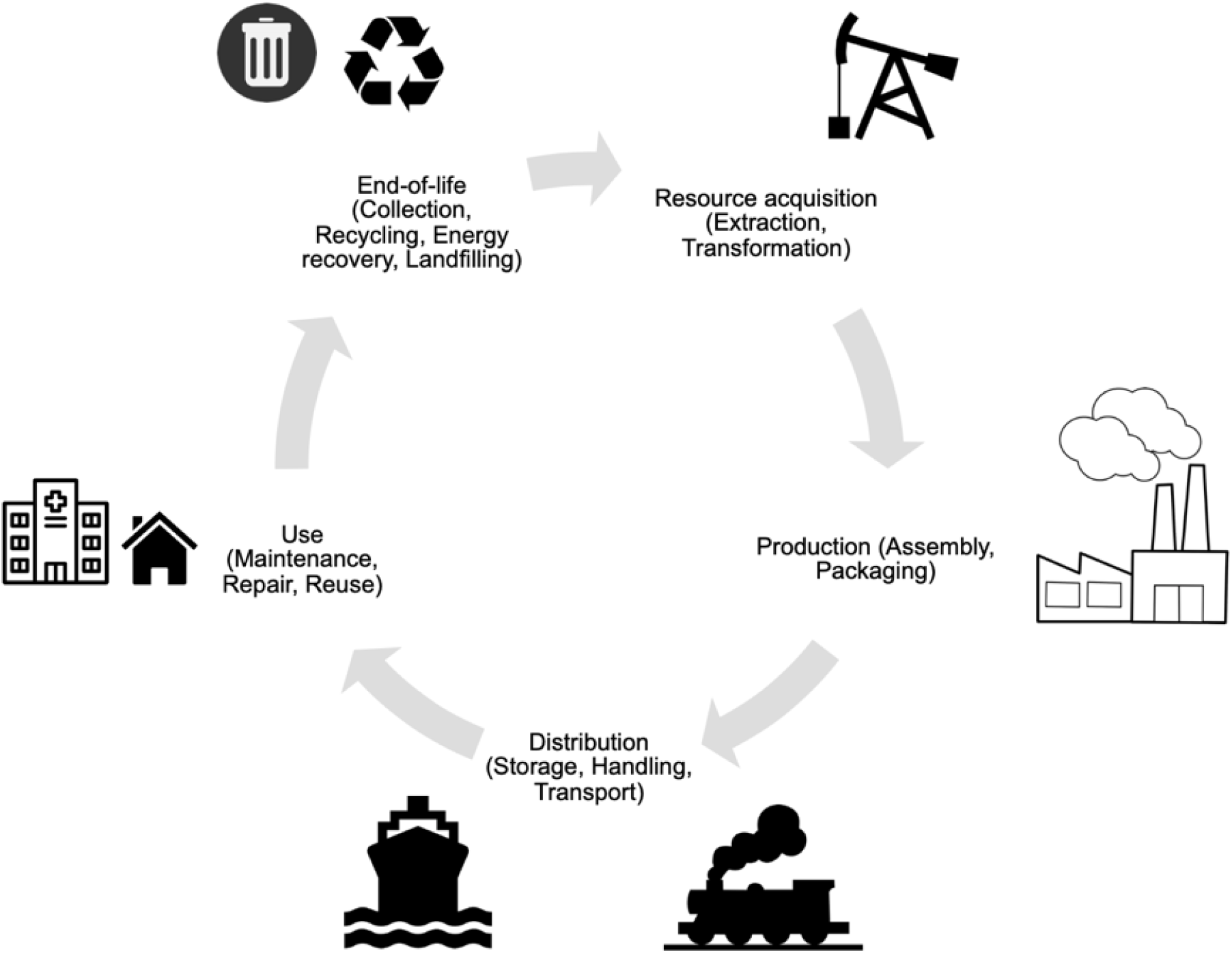
Components of life cycle assessment from cradle to grave. Figure printed with permission from Ms. Laurie Ouellet

### Life cycle assessment

We evaluated an MDI (100 µg of salbutamol/dose, 200 doses) and a nebulization ampoule (5 mg/mL, 10 mL), both products manufactured by GSK. An MDI is for single-patient use, whereas multiple patients can be treated with a single nebulization ampoule. The assessment followed ISO standards 14040 and 14044 ^9,10^ and consisted of 4 phases:

1 – Definition of the goal, scope, and functional unit We compared 1 and 3 administrations of a standard dose of salbutamol in the ED for adults and children weighing ≥ 24 kg, namely 800 *µ*g (8 puffs) via MDI and 5 mg (1 ml) via nebulization (functional unit). The life cycle assessment included production, packaging, transportation, use in the ED, and incineration, assuming all material disposal takes place at the hospital. We considered MDI treatments with a reusable plastic spacer or a single-use cardboard spacer, as well as all the reusable material (nebulizing cup, plastic tubing, oxygen mask) required for nebulization.
2 – Life cycle inventory We decomposed the life cycles of the 2 therapeutic options to identify all emissions and extractions within the system boundaries. In addition to the product monographs, we obtained the inventory data for all components of the MDI, plastic and cardboard spacers, nebulization ampoule and materials using attenuated total reflectance infrared spectroscopy, which provides detailed information about the molecular composition of a product. The composition of the MDI canister gas was analyzed using multinuclear nuclear magnetic resonance and gas-phase infrared spectroscopies. The methods for identifying the volatile components are described in e-Appendix 1. The production plants were located using procurement information obtained from our institution. The road distance to the biomedical waste management company used by the CHU (Stericycle) was determined. The complete life cycles were modeled using the openLCA software and the ecoinvent life cycle inventory database (version 3.9.1) and adapting some processes to the [name of the region] context.
3– Impact assessment and outcome measures The inventoried emissions and extractions were translated into environmental indicators in the following impact categories: i) *Climate change*, in kg of CO_2_ equivalent emissions (CO_2_eq); ii) *Human health*, in disability-adjusted life-year (DALY) loss; iii) *Fossil resources use*, in megajoules (MJ) deprived; and iv) *Ecosystem quality*, in potentially disappeared fraction of species (PDF) over square meter years (PDF ·m^2^·year). The human health and ecosystem quality indicators aggregate a series of intermediate effects of the production, use, and disposal of asthma treatment products. The inventory data were converted into impact units using the validated assessment method IPCC2021 GWP100 for *Climate change* and the IMPACT World+ method for the other impact categories.
4– Interpretation of results The indicator results were interpreted relative to the functional units for 1 or 3 administrations, which are common initial prescriptions for asthma treatment in many ED protocols. Hotspots were identified, that is, the life cycle elements having the highest contribution to the indicator.

### Alternative scenarios and extrapolations

Patients discharged from the CHU EDs are often allowed to keep the MDI for use at home. We considered this alternative scenario in the model by assuming that all such MDIs were emptied and ended up in the same incinerator as if they had been left in the ED.

We also estimated the annual environmental footprint of the asthma treatments based on medication consumption recorded at the CHU during 2022–2023. Since it was not possible to retrace the number of administrations and the dosages for each patient treated with an MDI, we assumed a minimum of 3 administrations of 800 *µ*g (8 inhalations) for each MDI delivered by the pharmacy to the ED during the study period.

### Statistical analysis

We report the results as point estimates and stacked bar charts in accordance with the standards used for life cycle assessment reporting. To account for the uncertainties of the proxies used in the inventory, we conducted a Monte Carlo simulation analysis with 1,000 iterations, from which we calculated point estimates for each indicator, differences between salbutamol administration modes, and 95% confidence intervals. All analyses were performed using openLCA (GreenDelta, version 2.0.4).

## RESULTS

### Life cycle inventories

The life cycle inventories of salbutamol treatments using MDIs and nebulization are reported in Table 1. The HFA used in the inhaler is 1,1,1,2-tetrafluoroethane, a gas having a global warming potential 1,500 times that of CO_2_.

**TABLE 1.**
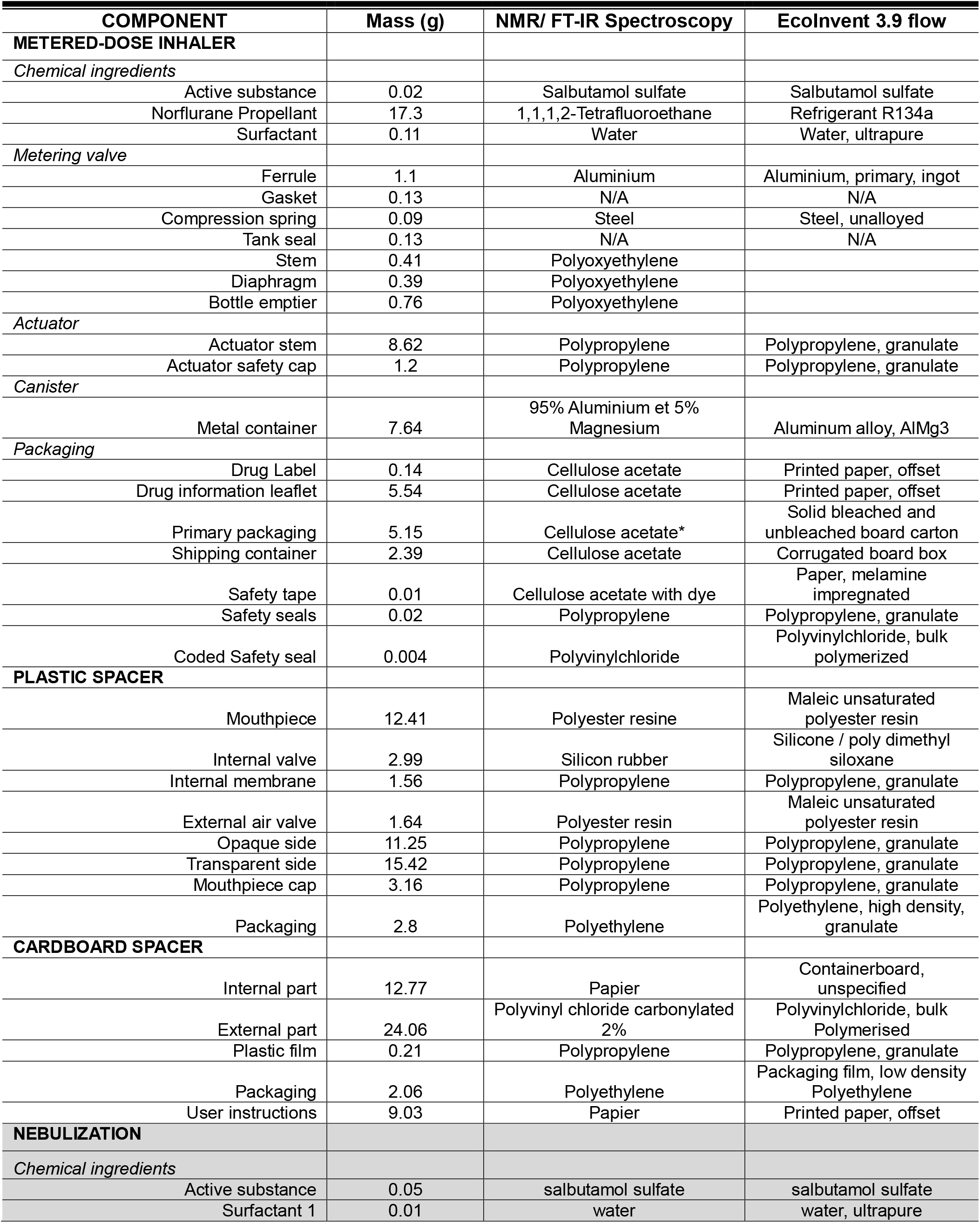
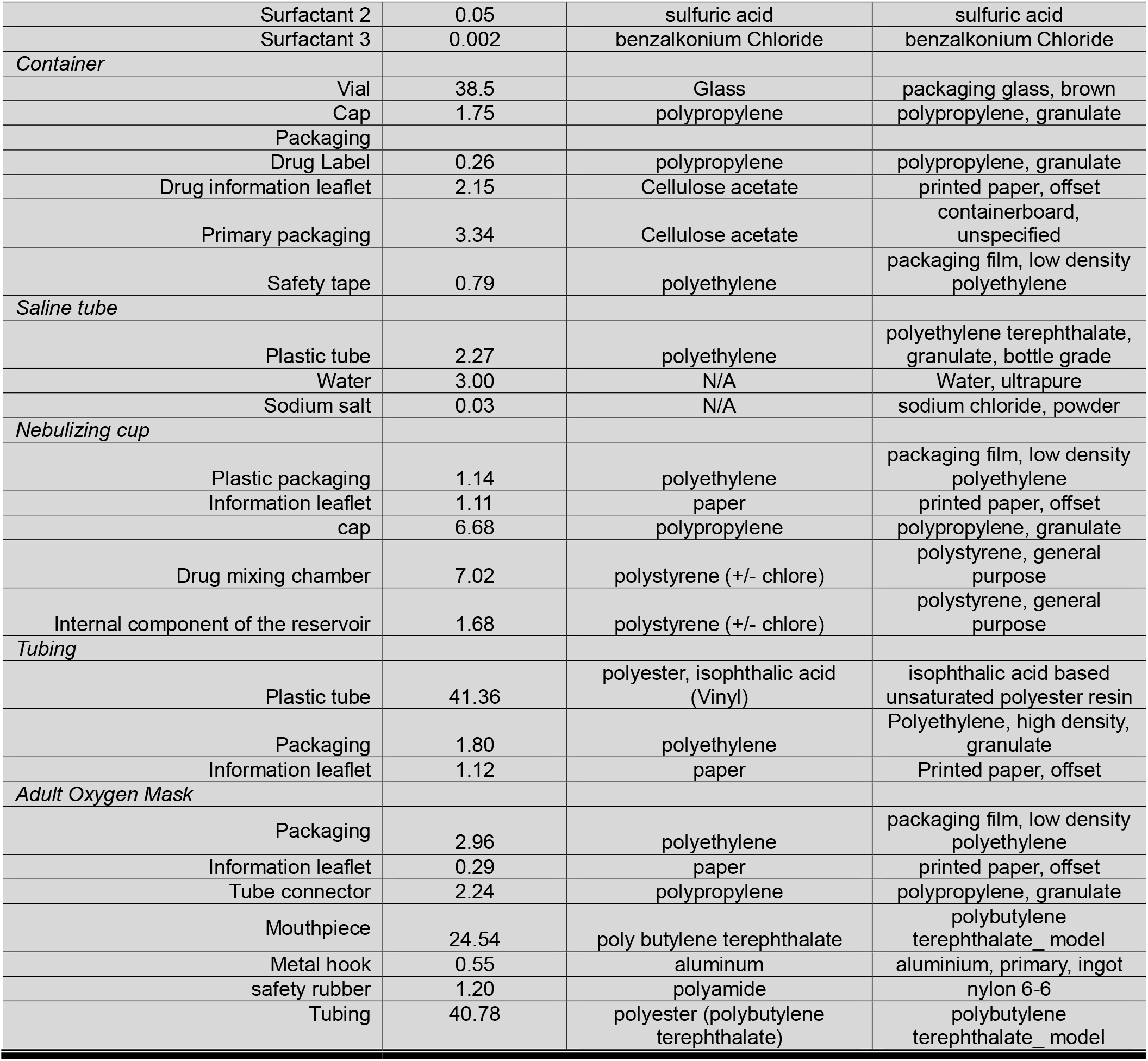
Life cycle inventories of salbutamol administration in emergency departments using metered-dose inhalers and nebulization.

| COMPONENT | Mass (g) | NMR/ FT-IR Spectroscopy | EcolInvent 3.9 flow |
| --- | --- | --- | --- |
| **METERED-DOSE INHALER** |  |  |  |
| *Chemical ingredients* |  |  |  |
| Active substance | 0.02 | Salbutamol sulfate | Salbutamol sulfate |
| Norflurane Propellant | 17.3 | 1,1,1,2-Tetrafluoroethane | Refrigerant R134a |
| Surfactant | 0.11 | Water | Water, ultrapure |
| *Metering valve* |  |  |  |
| Ferrule | 1.1 | Aluminium | Aluminium, primary, ingot |
| Gasket | 0.13 | N/A | N/A |
| Compression spring | 0.09 | Steel | Steel, unalloyed |
| Tank seal | 0.13 | N/A | N/A |
| Stem | 0.41 | Polyoxyethylene |  |
| Diaphragm | 0.39 | Polyoxyethylene |  |
| Bottle emptier | 0.76 | Polyoxyethylene |  |
| *Actuator* |  |  |  |
| Actuator stem | 8.62 | Polypropylene | Polypropylene, granulate |
| Actuator safety cap | 1.2 | Polypropylene | Polypropylene, granulate |
| *Canister* |  |  |  |
| Metal container | 7.64 | 95% Aluminium et 5% Magnesium | Aluminum alloy, AlMg3 |
| *Packaging* |  |  |  |
| Drug Label | 0.14 | Cellulose acetate | Printed paper, offset |
| Drug information leaflet | 5.54 | Cellulose acetate | Printed paper, offset |
| Primary packaging | 5.15 | Cellulose acetate* | Solid bleached and unbleached board carton |
| Shipping container | 2.39 | Cellulose acetate | Corrugated board box |
| Safety tape | 0.01 | Cellulose acetate with dye | Paper, melamine impregnated |
| Safety seals | 0.02 | Polypropylene | Polypropylene, granulate |
| Coded Safety seal | 0.004 | Polyvinylchloride | Polyvinylchloride, bulk polymerized |
| **PLASTIC SPACER** |  |  |  |
| Mouthpiece | 12.41 | Polyester resine | Maleic unsaturated polyester resin |
| Internal valve | 2.99 | Silicon rubber | Silicone / poly dimethyl siloxane |
| Internal membrane | 1.56 | Polypropylene | Polypropylene, granulate |
| External air valve | 1.64 | Polyester resin | Maleic unsaturated polyester resin |
| Opaque side | 11.25 | Polypropylene | Polypropylene, granulate |
| Transparent side | 15.42 | Polypropylene | Polypropylene, granulate |
| Mouthpiece cap | 3.16 | Polypropylene | Polypropylene, granulate |
| Packaging | 2.8 | Polyethylene | Polyethylene, high density, granulate |
| **CARDBOARD SPACER** |  |  |  |
| Internal part | 12.77 | Papier | Containerboard, unspecified |
| External part | 24.06 | Polyvinyl chloride carbonylated 2% | Polyvinylchloride, bulk Polymerised |
| Plastic film | 0.21 | Polypropylene | Polypropylene, granulate |
| Packaging | 2.06 | Polyethylene | Packaging film, low density Polyethylene |
| User instructions | 9.03 | Papier | Printed paper, offset |
| **NEBULIZATION** |  |  |  |
| *Chemical ingredients* |  |  |  |
| Active substance | 0.05 | salbutamol sulfate | salbutamol sulfate |
| Surfactant 1 | 0.01 | water | water, ultrapure |
| Surfactant 2 | 0.05 | sulfuric acid | sulfuric acid |
| Surfactant 3 | 0.002 | benzalkonium Chloride | benzalkonium Chloride |
| *Container* |  |  |  |
| Vial | 38.5 | Glass | packaging glass, brown |
| Cap | 1.75 | polypropylene | polypropylene, granulate |
| Packaging |  |  |  |
| Drug Label | 0.26 | polypropylene | polypropylene, granulate |
| Drug information leaflet | 2.15 | Cellulose acetate | printed paper, offset |
| Primary packaging | 3.34 | Cellulose acetate | containerboard, unspecified |
| Safety tape | 0.79 | polyethylene | packaging film, low density polyethylene |
| *Saline tube* |  |  |  |
| Plastic tube | 2.27 | polyethylene | polyethylene terephthalate, granulate, bottle grade |
| Water | 3.00 | N/A | Water, ultrapure |
| Sodium salt | 0.03 | N/A | sodium chloride, powder |
| *Nebulizing cup* |  |  |  |
| Plastic packaging | 1.14 | polyethylene | packaging film, low density polyethylene |
| Information leaflet | 1.11 | paper | printed paper, offset |
| cap | 6.68 | polypropylene | polypropylene, granulate |
| Drug mixing chamber | 7.02 | polystyrene (+/- chlore) | polystyrene, general purpose |
| Internal component of the reservoir | 1.68 | polystyrene (+/- chlore) | polystyrene, general purpose |
| *Tubing* |  |  |  |
| Plastic tube | 41.36 | polyester, isophthalic acid (Vinyl) | isophthalic acid based unsaturated polyester resin |
| Packaging | 1.80 | polyethylene | Polyethylene, high density, granulate |
| Information leaflet | 1.12 | paper | Printed paper, offset |
| *Adult Oxygen Mask* |  |  |  |
| Packaging | 2.96 | polyethylene | packaging film, low density polyethylene |
| Information leaflet | 0.29 | paper | printed paper, offset |
| Tube connector | 2.24 | polypropylene | polypropylene, granulate |
| Mouthpiece | 24.54 | poly butylene terephthalate | polybutylene terephthalate_model |
| Metal hook | 0.55 | aluminum | aluminium, primary, ingot |
| safety rubber | 1.20 | polyamide | nylon 6-6 |
| Tubing | 40.78 | polyester (polybutylene terephthalate) | polybutylene terephthalate_model |

The distances of the production plant and the incinerator (for both salbutamol administration methods) from the CHU were respectively 1605 km and 1813 km.

### Impact categories

The environmental footprint of the two methods is reported per impact category for 1 and 3 administrations in Tables 2 and 3, and e-Figures 1 to 4. The *Climate change* indicator hotspot for MDIs is HFA emission. Their carbon footprint is thus linked to their use: each series of 8 inhalations emits 1.1 kg of CO_2_eq as 1,1,1,2-tetrafluoroethane. This emission is also a notable contributor to the other indicators (especially in the case of 3 series) but MDI and spacer production and disposal (of the plastic spacer more than the cardboard) are also contributors. For nebulization, cup and mask production and disposal are hotspots for all indicators. Since these devices are reusable, indicator results do not increase much with the number of administrations. The results also indicate that although the single-use cardboard spacer leaves a smaller footprint than the plastic spacer after 1 administration, this difference diminishes or reverses in favor of the plastic spacer after 3 administrations.

**TABLE 2.** Climate change and human health indicators for salbutamol administration in the emergency department.

| # admin | Mode of administration | Climate change (kg CO<sub>2</sub> eq) |  | Human health (DALY) |  |
| --- | --- | --- | --- | --- | --- |
|  |  | Point estimate * | Median (IQR)** | Point estimate X 10<sup>-6</sup> | Median (IQR) X 10<sup>-6</sup> |
| 1 | MDI/plastic spacer | 1.89 | 1.91 (1.80; 2.05) | 4.1 | 1.7 (-64.3; 70.0) |
|  | MDI/cardboard spacer | 1.66 | 1.70 (1.58; 1.83) | 3.3 | 2.1 (-97.8; 99.3) |
|  | Nebulization | 0.95 | 1.10 (1.06; 1.14) | 3.9 | 13.0 (-191.2; 185.5) |
| 3 | MDI/plastic spacer | 4.00 | 4.05 (3.69; 4.43) | 6.0 | 5.8 (-55.6; 74.9) |
|  | MDI/cardboard spacer | 3.96 | 4.00 (3.66; 4.38) | 6.0 | 5.7 (-182.0; 193.7) |
|  | Nebulization | 0.99 | 1.13 (1.10; 1.18) | 4.0 | 2.7 (-186.0; 206.1) |

\*Point estimates obtained from impact evaluation of the life cycle assessment methodology
\*\*Medians and interquartile ranges (IQR) estimated using a Monte Carlo simulation (1,000 iterations)
#admin: number of administrations in the emergency department
MDI: metered-dose inhaler; CO<sub>2</sub>eq: CO<sub>2</sub> equivalent emissions; DALY: disability-adjusted life-year.

**TABLE 3.**
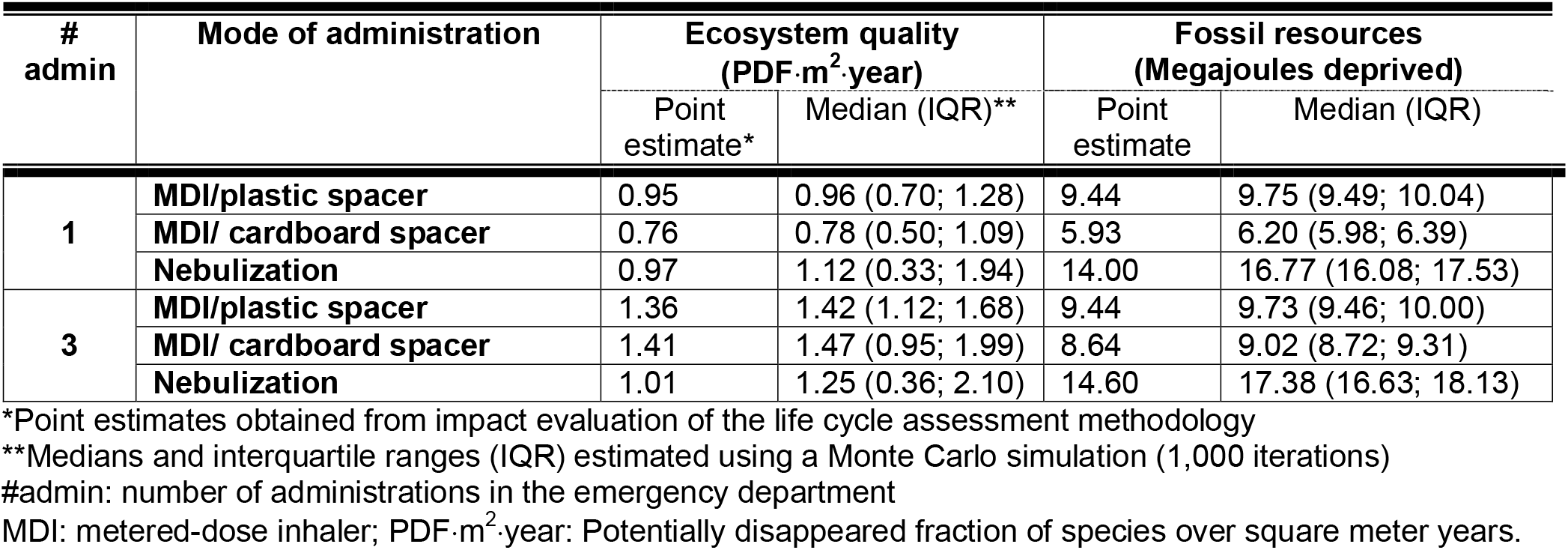
Biodiversity and fossil resource use indicators for salbutamol administration in the emergency department.

| # admin | Mode of administration | Ecosystem quality (PDF·m<sup>2</sup>·year) |  | Fossil resources (Megajoules deprived) |  |
| --- | --- | --- | --- | --- | --- |
|  |  | Point estimate* | Median (IQR)** | Point estimate | Median (IQR) |
| 1 | MDI/plastic spacer | 0.95 | 0.96 (0.70; 1.28) | 9.44 | 9.75 (9.49; 10.04) |
|  | MDI/ cardboard spacer | 0.76 | 0.78 (0.50; 1.09) | 5.93 | 6.20 (5.98; 6.39) |
|  | Nebulization | 0.97 | 1.12 (0.33; 1.94) | 14.00 | 16.77 (16.08; 17.53) |
| 3 | MDI/plastic spacer | 1.36 | 1.42 (1.12; 1.68) | 9.44 | 9.73 (9.46; 10.00) |
|  | MDI/ cardboard spacer | 1.41 | 1.47 (0.95; 1.99) | 8.64 | 9.02 (8.72; 9.31) |
|  | Nebulization | 1.01 | 1.25 (0.36; 2.10) | 14.60 | 17.38 (16.63; 18.13) |

\*Point estimates obtained from impact evaluation of the life cycle assessment methodology
\*\*Medians and interquartile ranges (IQR) estimated using a Monte Carlo simulation (1,000 iterations)
#admin: number of administrations in the emergency department
MDI: metered-dose inhaler; PDF·m<sup>2</sup>·year: Potentially disappeared fraction of species over square meter years.

Table 4 displays the median and 95% confidence interval of the difference between MDI and nebulization for each impact category. The *Climate change* impact is significantly greater for MDI than for nebulization, and this difference is greater for 3 administrations, reaching up to 2.9 kg CO_2_eq. The two salbutamol treatment methods are indistinguishable in the *Human health* category for 1 and 3 administrations. Nebulization has a greater negative impact on ecosystem quality after 1 administration, but the difference between the two methods is reversed after 3 administrations, as MDIs result in a greater loss of biodiversity. Nebulizing shows significantly higher impact on *Fossil resources use* than MDI at both 1 and 3 administrations.

**TABLE 4.** Differences between administering salbutamol via metered-dose inhaler and nebulization in the emergency department: environmental indicators.

| #admin | Modes | Climate change<br>(kg CO<sub>2</sub> eq) | Human health<br>(DALY)<br>X 10<sup>-5</sup> | Ecosystem quality<br>(PDF·m<sup>2</sup>·year) | Fossil resources<br>(MJ deprived) |
| --- | --- | --- | --- | --- | --- |
| 1 | MDI/plastic spacer<br>minus Nebul | 0.85<br>(0.84 to 0.86)* | 0.84<br>(-0.31 to 1.99) | -0.12<br>(-0.17 to -0.07) | -6.99<br>(-7.06 to -6.92) |
|  | MDI/cardboard<br>spacer minus Nebul | 0.61<br>(0.60 to 0.62) | -1.05<br>(-2.37 to 0.26) | -0.37<br>(-0.43 to -0.32) | -10.58<br>(-10.65 to -10.51) |
| 3 | MDI/plastic spacer<br>minus Nebul | 2.92<br>(2.89 to 2.96) | -0.93<br>(-2.10 to 0.25) | 0.21<br>(0.15 to 0.26) | -7.68<br>(-7.75 to -7.61) |
|  | MDI/cardboard<br>spacer minus Nebul | 2.91<br>(2.88 to 2.95) | -0.20<br>(-1.85 to 1.45) | 0.30<br>(0.25 to 0.35) | -8.38<br>(-8.45 to -8.31) |

\*Difference (95% Confidence Interval) between the treatment options estimated using a Monte Carlo simulation (1,000 iterations)
#admin: number of administrations
MDI: metered-dose inhaler; CO<sub>2</sub>eq: CO<sub>2</sub> equivalent emissions;
DALY: disability-adjusted life-year;

### Metered-dose inhaler left to the patient for home use

The environmental footprint of the scenario in which the patient upon discharge is allowed to keep the MDI for use at home is reported in e-Table 1. For all categories, the impact of this scenario is lower than disposal of the MDI at the hospital. While the contribution of HFA emission continues to increase, the contribution of MDI production and disposal is spread over 200 inhalations instead of 8 or 24 (1 or 3 administrations) per functional unit. The previously observed differences between MDIs and nebulization for all indicators do not change significantly under this scenario. We estimated a difference of 2.5–2.6 kg CO_2_eq between MDI and nebulization for 3 treatments, with no significant differences noted in other categories of impact.

### One-year ecological footprint

In fiscal year 2022–2023, 4,815 MDIs and 233 nebulizer ampoules were dispensed in the 5 EDs associated with the CHU. The corresponding annual environmental footprint is reported by impact category in Table 5, based on each patient receiving 3 administrations of salbutamol while in the ED. An emission of 19,260 kg of CO_2_ equivalent is attributable to the use of MDIs. If all patients had received nebulization, this emission would have been 14,416 kg lower, and losses of 0.93 × 10^−2^ DALY and 1,606 PDF·m^2^·year would have been avoided, but an additional 25,984 MJ in fossil resources would have been consumed. The 14,416 kg of CO_2_eq emissions is equivalent to driving 84,800 kilometers in a subcompact car or 19 trips from New York to Los Angeles.

**TABLE 5.**
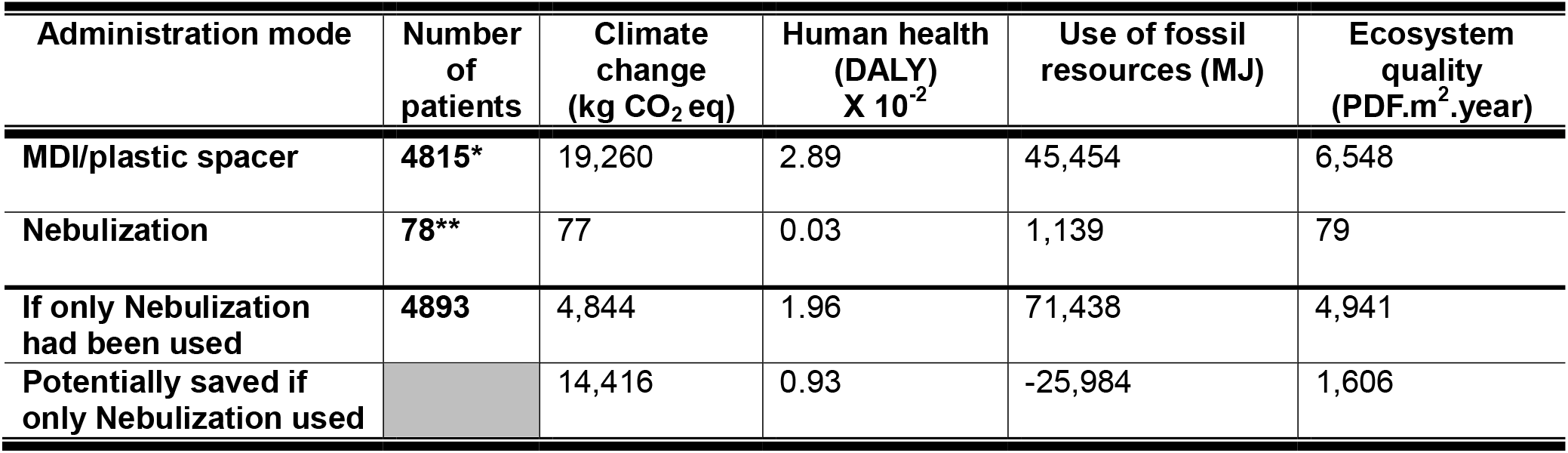
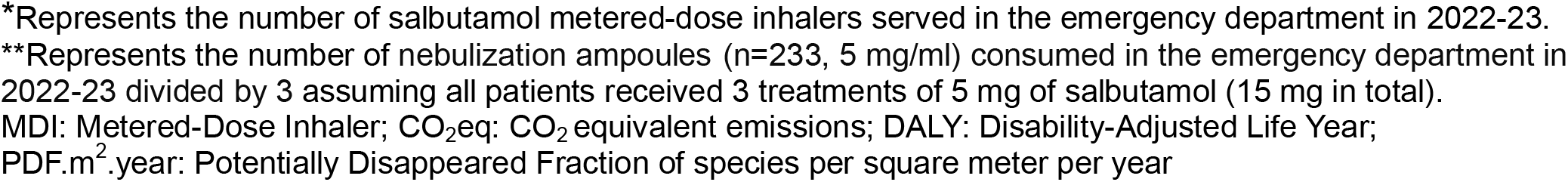
Extrapolation of the Ecological Footprint of Salbutamol Administration via Metered-Dose Inhaler and Nebulization According to Medication Consumption Recorded at the CHU in Fiscal Year 2022-23.

| Administration mode | Number of patients | Climate change (kg CO<sub>2</sub> eq) | Human health (DALY) X 10<sup>-2</sup> | Use of fossil resources (MJ) | Ecosystem quality (PDF·m<sup>2</sup>·year) |
| --- | --- | --- | --- | --- | --- |
| MDI/plastic spacer | 4815* | 19,260 | 2.89 | 45,454 | 6,548 |
| Nebulization | 78** | 77 | 0.03 | 1,139 | 79 |
| If only Nebulization had been used | 4893 | 4,844 | 1.96 | 71,438 | 4,941 |
| Potentially saved if only Nebulization used |  | 14,416 | 0.93 | -25,984 | 1,606 |

\*Represents the number of salbutamol metered-dose inhalers served in the emergency department in 2022-23. \*\*Represents the number of nebulization ampoules (n=233, 5 mg/ml) consumed in the emergency department in 2022-23 divided by 3 assuming all patients received 3 treatments of 5 mg of salbutamol (15 mg in total). MDI: Metered-Dose Inhaler; CO<sub>2</sub>eq: CO<sub>2</sub> equivalent emissions; DALY: Disability-Adjusted Life Year; PDF.m<sup>2</sup>.year: Potentially Disappeared Fraction of species per square meter per year

## DISCUSSION

In this product/process life cycle assessment, we examined ED treatment of acute asthma using salbutamol administration via MDIs versus nebulization. Nebulization was found to have half the carbon footprint of 1 MDI administration and one quarter of 3 MDI administrations. The two methods are indistinguishable in the *Human health* category for 1 and 3 administrations, and in the *Ecosystem quality* category for 1 administration but not 3, at which the MDI had a significantly higher impact. Nebulizing shows significantly higher impact on *Fossil resources use* for 1 or 3 MDI administrations. When scaled to the 5,000 patients treated annually for asthma in the EDs of the institution where this research was conducted, nearly 20,000 kg CO_2_ equivalent emissions would be attributable to the use of MDIs and nearly 15,000 kg would be avoided by prescribing nebulization.

### Previous literature

To our knowledge, this is the first study comparing the environmental footprint of MDIs and nebulization in the ED. Goulet et al. compared HFA-based MDIs to a portable nebulizer device for use at home by patients with COPD ^11^. Comparisons with their study should be made carefully since: 1) a different functional unit was used (200 *µ*g of Albuterol by MDI and 3 mg by nebulizer); 2) the nebulizer device studied is not used in the ED; and 3) MDI administration was modeled alone without a spacer. Still, the authors calculated a carbon footprint 2 to 3 times higher for one dose with an MDI compared to one dose with a nebulizer, whereas we estimated that one emergency treatment with an MDI has twice the carbon footprint of nebulization. In absolute numbers, their CO_2_eq associated with MDI use appears disproportionately low (0.097 kg CO_2_eq for 2 inhalations) compared to our estimates (1.89 kg CO_2_eq for 8 inhalations). However, the Goulet et al. model contained 6.7 g of HFA compared to 17.3 g used in our calculations. Their method of estimating the quantity of HFA is not described in detail, whereas our approach is reproducible and aligns with the quantities estimated another study that focused on gas kinetics associated with pressurized MDIs ^12^.

We also modeled the scenario in which the patient uses the MDI at home, which was found to lead to a lower carbon footprint. However, dry powder inhalers (DPI), which can be used by patients aged 6 and above ^13^, have been found comparable in efficacy ^14^ and cost ^15^ to MDIs and to have a carbon footprint 20-30 times smaller ^16^. If DPIs are in fact usable at home in the same manner as bronchodilators, then discarding MDIs after use in the ED would become the approach having the least impact as a source of greenhouse gas emission.

### Future direction

This study of a specific aspect of the environmental footprint of healthcare systems suggests that nebulization is a more eco-efficient method of administering bronchodilators in EDs that receive many patients needing such treatments. However, particularly in children, this method has been associated with more side effects and longer ED stays ^8^. Further research is needed to determine if the use of DPIs is effective and safe as ED treatment of mild to moderate acute asthma, which would allow nebulization to be reserved for the most severe cases.

### Limitations

Among the limitations of this study, we must mention our creation of the life cycle inventories without access to industry information. However, the method used is scientifically robust and reproducible, and we are confident that the resulting analysis adequately estimates the differences in environmental footprint between MDI use and nebulization. Our results might have differed if other inventory databases and impact assessment methods had been used. However, the ecoinvent database is one of the most widely used in the world, and processes were adapted to the [name of the region] context when this was relevant. The CO_2_eq assessment method is that proposed by the Intergovernmental Panel on Climate Change, a widely recognized international organization created under the auspices of the UN to assess risks related to climate change. We believe that this choice of tools makes our analysis as robust and credible as presently possible. Finally, to estimate the one-year environmental footprint, we assumed that patients received their 3 MDI treatments at 20-minute intervals, since this is the protocolized prescription for asthma treatment in the CHU emergency departments. Since a significant number of patients end up receiving more than 3 treatments, our extrapolations are likely underestimates of the real impact.

## CONCLUSION

Acute asthma is one of the most frequent conditions treated in emergency departments. The administration of salbutamol for this purpose usually involves the use of an MDI, which has a significant environmental footprint. Our study shows that administering salbutamol by nebulization has a smaller footprint. Given that more and more governments around the world are committed to providing healthcare services with net-zero greenhouse gas emissions over the coming decades, implementing low-emission ED treatment protocols for acute asthma should be one of many avenues to explore to achieve this goal.

## Supporting information

web-appendix A1-A6

## Data Availability

All data produced in the present study are available upon reasonable request to the authors.

## Abbreviations

ED: emergency department
CO_2_eq: CO_2_ equivalents
MDI: metered-dose inhalers
COPD: chronic obstructive pulmonary disease
HFA: hydrofluoroalkane
DALY: disability-adjusted life-year
MJ: megajoule
PDF m^2^ year: potentially disappeared fraction of species over square meter years
IPCC2021 GWP100: Intergovernmental Panel on Climate Change for 2021 – Global Warming Potential for 100-year period
DPI: dry powder inhaler

## Aknowledgement

The authors thank Ms. Laurie Ouellet for granting permission to use her figure illustrating the life cycle.

