## Supplementary material for "Comparison of the Ecological Footprints of administering Salbutamol by Metered-Dose Inhaler and by Nebulization in Emergency Treatment of Acute Asthma": web-appendix A1-A6

**WEB-APPENDICES**

**e-Appendix 1. Description of the methods for identifying gas composition in the metered-dose inhaler canister**

The identity of the major volatile components of the mixture within salbutamol canisters was ascertained by multinuclear nuclear magnetic resonance (NMR) spectroscopy and gas-phase infrared (IR) spectroscopy. Volatile but condensable (most volatiles excluding extremes like N_2_, O_2_, etc.) substances were collected using a setup inspired by ref^1^ consisting of an anesthesia bag, into which the salbutamol canister and pump assembly was placed, connected to a high-vacuum line with calibrated volumes equipped with a train of U-traps for fractional condensations. Such a setup allowed for the exclusion of moisture from the environment. Ideal gas behavior was used as an approximation and the ideal gas law was used to determine the amount of materials collected in the gas phase. This approximation is widely useable, especially with species well above their boiling point, and typically gives results well within 95 % accuracy when compared with weighing with an analytical balance.

^1^H, ^19^F and ^13^C NMR supported by MS indicated that the mixture contained CF_3_CFH_2_ as the “HFA” propellant.

A canister of GSK brand salbutamol was weighed before (28.7140 g) and after (11.4611 g) being emptied by 200-300 actuations of the pump (17.2582 g of material expelled) The last portion, consisting mostly of gas-phase material, was particularly difficult to extract. We attempted to disassemble the canister by essentially ripping the top apart while cooling the canister to -196 °C (Caution! This operation is dangerous and should only be attempted with proper safety equipment and gas-phase handling techniques). The open canister was quickly placed in a vacuum vessel and attached to high-vacuum line. The volatiles were then collected (empty canister + disassembled parts : 10.9986 g) and approx. 0.3 mmol of material were collected (*maximum 31 mg if composed of CF_3_CH_2_F)*, yielding an estimated total mass of volatile components (excluding the non-volatile salbutamol) of between 17.26 g and 17.7 g. This is in agreement with ref^1^, which found *ca* 17.32 g of propellant (likely excluding the gas-phase remainder in the canister).

The composition of the mixture was assumed to be uniform throughout actuations because of the design of the pump, which shoots out the suspension (comprising the liquid propellant, solid salbutamol and other potential formulation ingredients) that vaporizes upon exiting the canister. A sample consisting of the 15 first actuations (1.1204 g weight loss) of a new salbutamol canister (GSK brand) was collected by condensation at -196 °C (1.085 g total based on ideal gas law). Moisture (most likely adventitiously condensed along the propellant during manufacture) could be visually observed in the process in the form of a white, high-melting point solid condensed in the traps. This solid mixture was then fractionally condensed through U-traps at -96 °C, -130 °C and -196 °C under a dynamic vacuum. Approximately (80 μmol, 8 mg)) out of the total volatile components were separated and identified by ^1^H NMR spectroscopy and vibrational spectroscopy as water. The rest of the major volatile components were confirmed to be tetrafluoroethane with possible traces of moisture (1.07 g based on ideal gas law). The composition was therefore found to be in agreement with the literature provided with the pump, which lists tetrafluoroethane as the only non-medicinal ingredient.

The literature^1^ indicates the use of ethanol in salbutamol pumps. Indeed, a sample of Teva brand salbutamol was found to contain ethanol, as indicated by ^1^H NMR spectroscopy. ^1^H NMR relative signal integrations, which are a crude approximation for the true composition, suggest a proportion of at most 5 mol% ethanol (2 w/w%). Similarly, semi-quantitative MS data also suggests a concentration of *ca* 5 mol% of ethanol. These results are in qualitative agreement with ref^1^ (1 w/w % ethanol). Despite our best efforts, no ethanol was detected in samples of the GSK brand salbutamol, in agreement with its listed non-medicinal ingredients, which omit ethanol. CO_2_ (most likely adventitiously incorporated during the manufacture) was also identified by ^13^C spectroscopy in samples of the Teva brand salbutamol.

[1] W. F. S. Sellers, *Allergy, Asthma & Clinical Immunology* **2017**, *13*, 30.

e-Figure 1. CO2 equivalent emissions by Emergency Department treatment of asthma with salbutamol, calculated using the IPCC2021 GWP100 method

kg CO2eq

|  | MDI use (HFA) | MDI production | Salbutamol production | Spacer production | MDI end-of-life | Spacer end-of-life | HFA end-of-life | Salbutamol end-of-life | H2O end-of-life | Vial production | Salbutamol production | Cup production | Mask production | Saline vial production | Vial end-of-life | Cup end-of-life | Mask end-of-life | Saline vial end-of-life |
| --- | --- | --- | --- | --- | --- | --- | --- | --- | --- | --- | --- | --- | --- | --- | --- | --- | --- | --- |
| MDI - Plastic spacer 1 admin, discarded in the ED | 1.05619 | 0.444144 | 0.000716445 | 0.186669 | 0.0508298 | 0.129268 | 0.0191655 | 4.347e-05 | 0.000285511 |  |  |  |  |  |  |  |  |  |
| MDI - Plastic spacer 3 admins, discarded in the ED | 3.16857 | 0.444144 | 0.000716445 | 0.186669 | 0.0508298 | 0.129268 | 0.0175683 | 3.98475e-05 | 0.000261719 |  |  |  |  |  |  |  |  |  |
| MDI - Plastic spacer 1 admin, emptied by patient | 1.05619 | 0.0177658 | 2.86578e-05 | 0.186669 | 0.00203319 | 0.129268 | 0 | 0 | 0 |  |  |  |  |  |  |  |  |  |
| MDI - Plastic spacer 3 admins, emptied by patient | 3.16857 | 0.0532973 | 8.59733e-05 | 0.186669 | 0.00609958 | 0.129268 | 0 | 0 | 0 |  |  |  |  |  |  |  |  |  |
| MDI - Cardboard spacer 1 admin, discarded in the ED | 1.05619 | 0.444144 | 0.000716445 | 0.0820545 | 0.0508298 | 0.0105782 | 0.0191655 | 4.347e-05 | 0.000285511 |  |  |  |  |  |  |  |  |  |
| MDI - Cardboard spacer 3 admins, discarded in the ED | 3.16857 | 0.444144 | 0.000716445 | 0.246163 | 0.0508298 | 0.0317347 | 0.0175683 | 3.98475e-05 | 0.000261719 |  |  |  |  |  |  |  |  |  |
| MDI - Cardboard spacer 1 admin, emptied by patient | 1.05619 | 0.0177658 | 2.86578e-05 | 0.0820545 | 0.00203319 | 0.0105782 | 0 | 0 | 0 |  |  |  |  |  |  |  |  |  |
| MDI - Cardboard spacer 3 admins, emptied by patient | 3.16857 | 0.0532973 | 8.59733e-05 | 0.246163 | 0.00609958 | 0.0317347 | 0 | 0 | 0 |  |  |  |  |  |  |  |  |  |
| Nebulization 1 admin |  |  |  |  |  |  |  |  |  | 0.00584631 | 0.000179111 | 0.264553 | 0.322806 | 0.00676281 | 0.0007971 | 0.154719 | 0.155556 | 0.00478671 |
| Nebulization 3 admins |  |  |  |  |  |  |  |  |  | 0.0175389 | 0.000537333 | 0.264553 | 0.322806 | 0.0202884 | 0.0023913 | 0.154719 | 0.155556 | 0.0143601 |

IPCC2021 GWP100: Intergovernmental Panel on Climate Change 2021 Global Warming Potential over a 100-year period

CO_2_eq: CO2 equivalent emissions

Admin: Administration

ED: Emergency Department

MDI: Metered-dose inhaler

HFA: Hydrofluoroalkane

e-Figure 2. Disability-adjusted life-year loss attributable to the environmental impact of different modes of salbutamol administration in the Emergency Department

DALY

|  | MDI use (HFA) | MDI production | Salbutamol production | Spacer production | MDI end-of-life | Spacer end-of-life | HFA end-of-life | Salbutamol end-of-life | H2O end-of-life | Vial production | Salbutamol production | Cup production | Mask production | Saline vial production | Vial end-of-life | Cup end-of-life | Mask end-of-life | Saline vial end-of-life |
| --- | --- | --- | --- | --- | --- | --- | --- | --- | --- | --- | --- | --- | --- | --- | --- | --- | --- | --- |
| MDI - Plastic spacer 1 admin, discarded in the ED | 9.42323e-07 | 1.94605e-06 | 4.36565e-09 | 8.64153e-07 | 1.99234e-07 | 4.85543e-07 | 1.64597e-10 | 7.39337e-08 | 1.30233e-09 |  |  |  |  |  |  |  |  |  |
| MDI - Plastic spacer 3 admins, discarded in the ED | 2.82697e-06 | 1.94605e-06 | 4.36565e-09 | 8.64153e-07 | 1.99234e-07 | 4.85543e-07 | 1.50881e-10 | 6.77725e-08 | 1.1938e-09 |  |  |  |  |  |  |  |  |  |
| MDI - Plastic spacer 1 admin, emptied by patient | 9.42323e-07 | 7.78422e-08 | 1.74626e-10 | 8.64153e-07 | 7.96935e-09 | 4.85543e-07 | 0 | 0 | 0 |  |  |  |  |  |  |  |  |  |
| MDI - Plastic spacer 3 admins, emptied by patient | 2.82697e-06 | 2.33527e-07 | 5.23878e-10 | 8.64153e-07 | 2.3908e-08 | 4.85543e-07 | 0 | 0 | 0 |  |  |  |  |  |  |  |  |  |
| MDI - Cardboard spacer 1 admin, discarded in the ED | 9.42323e-07 | 1.94605e-06 | 4.36565e-09 | 4.83488e-07 | 1.99234e-07 | 4.45459e-08 | 1.64597e-10 | 7.39337e-08 | 1.30233e-09 |  |  |  |  |  |  |  |  |  |
| MDI - Cardboard spacer 3 admins, discarded in the ED | 2.82697e-06 | 1.94605e-06 | 4.36565e-09 | 4.83488e-07 | 1.99234e-07 | 4.45459e-08 | 1.50881e-10 | 6.77725e-08 | 1.1938e-09 |  |  |  |  |  |  |  |  |  |
| MDI - Cardboard spacer 1 admin, emptied by patient | 9.42323e-07 | 7.78422e-08 | 1.74626e-10 | 4.83488e-07 | 7.96935e-09 | 4.45459e-08 | 0 | 0 | 0 |  |  |  |  |  |  |  |  |  |
| MDI - Cardboard spacer 3 admins, emptied by patient | 2.82697e-06 | 2.33527e-07 | 5.23878e-10 | 4.83488e-07 | 2.3908e-08 | 4.45459e-08 | 0 | 0 | 0 |  |  |  |  |  |  |  |  |  |
| Nebulization 1 admin |  |  |  |  |  |  |  |  |  | 3.18716e-08 | 1.09141e-09 | 1.35302e-06 | 2.16145e-06 | 3.6419e-08 | 3.29247e-09 | 5.99495e-07 | 5.85664e-07 | 1.80118e-08 |
| Nebulization 3 admins |  |  |  |  |  |  |  |  |  | 9.56147e-08 | 3.27424e-09 | 1.35302e-06 | 2.16145e-06 | 1.09257e-07 | 9.8774e-09 | 5.99495e-07 | 5.85664e-07 | 5.40353e-08 |

DALY: Disability-Adjusted Life Year

MDI: Metered-dose inhaler

HFA: Hydrofluoroalkane

e-Figure 3. Potential loss of biodiversity attributable to different modes of salbutamol administration in the Emergency Department

PDF.m2.an

|  | MDI use (HFA) | MDI production | Salbutamol production | Spacer production | MDI end-of-life | Spacer end-of-life | HFA end-of-life | Salbutamol end-of-life | H2O end-of-life | Vial production | Salbutamol production | Cup production | Mask production | Saline vial production | Vial end-of-life | Cup end-of-life | Mask end-of-life | Saline vial end-of-life |
| --- | --- | --- | --- | --- | --- | --- | --- | --- | --- | --- | --- | --- | --- | --- | --- | --- | --- | --- |
| MDI - Plastic spacer 1 admin, discarded in the ED | 0.20592 | 0.367189 | 0.000832303 | 0.190833 | 0.0508208 | 0.126229 | 4.30803e-05 | 0.0193665 | 0.000323705 |  |  |  |  |  |  |  |  |  |
| MDI - Plastic spacer 3 admins, discarded in the ED | 0.61776 | 0.367189 | 0.000832303 | 0.190833 | 0.0508208 | 0.126229 | 3.94902e-05 | 0.0177527 | 0.000296729 |  |  |  |  |  |  |  |  |  |
| MDI - Plastic spacer 1 admin, emptied by patient | 0.20592 | 0.0146875 | 3.32921e-05 | 0.190833 | 0.00203283 | 0.126229 | 0 | 0 | 0 |  |  |  |  |  |  |  |  |  |
| MDI - Plastic spacer 3 admins, emptied by patient | 0.61776 | 0.0440626 | 9.98764e-05 | 0.190833 | 0.0060985 | 0.126229 | 0 | 0 | 0 |  |  |  |  |  |  |  |  |  |
| MDI - Cardboard spacer 1 admin, discarded in the ED | 0.20592 | 0.367189 | 0.000832303 | 0.115045 | 0.0508208 | 0.0104404 | 4.30803e-05 | 0.0193665 | 0.000323705 |  |  |  |  |  |  |  |  |  |
| MDI - Cardboard spacer 3 admins, discarded in the ED | 0.61776 | 0.367189 | 0.000832303 | 0.115045 | 0.0508208 | 0.0104404 | 3.94902e-05 | 0.0177527 | 0.000296729 |  |  |  |  |  |  |  |  |  |
| MDI - Cardboard spacer 1 admin, emptied by patient | 0.20592 | 0.0146875 | 3.32921e-05 | 0.115045 | 0.00203283 | 0.0104404 | 0 | 0 | 0 |  |  |  |  |  |  |  |  |  |
| MDI - Cardboard spacer 3 admins, emptied by patient | 0.61776 | 0.0440626 | 9.98764e-05 | 0.115045 | 0.0060985 | 0.0104404 | 0 | 0 | 0 |  |  |  |  |  |  |  |  |  |
| Nebulization 1 admin |  |  |  |  |  |  |  |  |  | 0.00745688 | 0.000208129 | 0.278055 | 0.343698 | 0.00699777 | 0.000824378 | 0.15087 | 0.151511 | 0.00466178 |
| Nebulization 3 admins |  |  |  |  |  |  |  |  |  | 0.0223707 | 0.000624388 | 0.278055 | 0.343698 | 0.0209933 | 0.00247313 | 0.15087 | 0.151511 | 0.0139853 |

PDF.m^2^.year: Potential Loss of Biodiversity per square meter per year

MDI: Metered-dose inhaler

HFA: Hydrofluoroalkane

e-Figure 4. Fossil energy resources use attributable to different modes of salbutamol administration in the Emergency Department

MJ deprived

|  | MDI use (HFA) | MDI production | Salbutamol production | Spacer production | MDI end-of-life | Spacer end-of-life | HFA end-of-life | Salbutamol end-of-life | H2O end-of-life | Vial production | Salbutamol production | Cup production | Mask production | Saline vial production | Vial end-of-life | Cup end-of-life | Mask end-of-life | Saline vial end-of-life |
| --- | --- | --- | --- | --- | --- | --- | --- | --- | --- | --- | --- | --- | --- | --- | --- | --- | --- | --- |
| MDI - Plastic spacer 1 admin, discarded in the ED | 0 | 4.33642 | 0.0115 | 4.82923 | 0.13922 | 0.04727 | 8.13423e-05 | 0.07397 | 0.00435 |  |  |  |  |  |  |  |  |  |
| MDI - Plastic spacer 3 admins, discarded in the ED | 0 | 4.33642 | 0.0115 | 4.82923 | 0.13922 | 0.04727 | 7.45638e-05 | 0.0678058 | 0.0039875 |  |  |  |  |  |  |  |  |  |
| MDI - Plastic spacer 1 admin, emptied by patient | 0 | 0.173457 | 0.00046 | 4.82923 | 0.0055688 | 0.04727 | 0 | 0 | 0 |  |  |  |  |  |  |  |  |  |
| MDI - Plastic spacer 3 admins, emptied by patient | 0 | 0.52037 | 0.00138 | 4.82923 | 0.0167064 | 0.04727 | 0 | 0 | 0 |  |  |  |  |  |  |  |  |  |
| MDI - Cardboard spacer 1 admin, discarded in the ED | 0 | 4.33642 | 0.0115 | 1.33598 | 0.13922 | 0.02235 | 8.13423e-05 | 0.07397 | 0.00435 |  |  |  |  |  |  |  |  |  |
| MDI - Cardboard spacer 3 admins, discarded in the ED | 0 | 4.33642 | 0.0115 | 1.33598 | 0.13922 | 0.02235 | 7.45638e-05 | 0.0678058 | 0.0039875 |  |  |  |  |  |  |  |  |  |
| MDI - Cardboard spacer 1 admin, emptied by patient | 0 | 0.173457 | 0.00046 | 1.33598 | 0.0055688 | 0.02235 | 0 | 0 | 0 |  |  |  |  |  |  |  |  |  |
| MDI - Cardboard spacer 3 admins, emptied by patient | 0 | 0.52037 | 0.00138 | 1.33598 | 0.0167064 | 0.02235 | 0 | 0 | 0 |  |  |  |  |  |  |  |  |  |
| Nebulization 1 admin |  |  |  |  |  |  |  |  |  | 0.09825 | 0.00288 | 6.07308 | 7.42531 | 0.17327 | 0.00456 | 0.06487 | 0.06803 | 0.00212 |
| Nebulization 3 admins |  |  |  |  |  |  |  |  |  | 0.29475 | 0.00864 | 6.07308 | 7.42531 | 0.51981 | 0.01368 | 0.06487 | 0.06803 | 0.00636 |

MDI: Metered-dose inhaler

HFA: Hydrofluoroalkane

MJ: Megajoules

**e-Table 1. Environmental impacts of Salbutamol administration modes for 1 and 3 administrations in the emergency department when metered-dose inhalers and spacers are left for patient’s use at home**

| **Number of times administered** | **Mode of administration** | **Climate change**  **(kg CO_2_ eq)** | | **Human health**  **(DALY)** | | **Ecosystem quality**  **(PDF⋅m^2^⋅year)** | | **Use of fossil resources**  **(Megajoules deprived)** | |
| --- | --- | --- | --- | --- | --- | --- | --- | --- | --- |
|  |  | Point estimate* | Monte Carlo**  Median (IQR) | Point estimate  X 10^-6^ | Monte Carlo  Median (IQR)  X 10^-6^ | Point estimate | Monte Carlo  Median (IQR) | Point estimate | Monte Carlo  Median (IQR) |
| **1** | **MDI/plastic spacer** | 1.39 | 1.42  (1.29; 1.54) | 2.3 | 0.7  (-25.7; 25.2) | 0.54 | 0.55  (0.43; 0.67) | 5.06 | 5.17  (5.00; 5.37) |
|  | **MDI/cardboard spacer** | 1.17 | 1.18  (1.06; 1.30) | 1.4 | -3.8  (-57.3; 51.3) | 0.34 | 0.36  (0.22; 0.50) | 1.54 | 1.61  (1.54; 1.68) |
|  | **Nebulization** | 0.95 | 1.10  (1.06; 1.14) | 3.9 | 13.0  (-191.2; 185.5) | 0.97 | 1.12  (0.33; 1.94) | 14.00 | 16.77  (16.08; 17.53) |
| **3** | **MDI/plastic spacer** | 3.54 | 3.57  (3.23; 3.90) | 4.3 | 6.1  (-19.9; 34.8) | 0.98 | 1.00  (0.86; 1.15) | 5.42 | 5.54  (5.34; 5.73) |
|  | **MDI/cardboard spacer** | 3.51 | 3.53  (3.16; 3.95) | 4.3 | 18.7  (-131.3; 177.5) | 1.03 | 1.08  (0.61; 1.47) | 4.61 | 4.82  (4.60; 5.06) |
|  | **Nebulization** | 0.99 | 1.13  (1.10; 1.18) | 4.0 | 2.7  (-186.0; 206.1) | 1.01 | 1.25  (0.36; 2.10) | 14.60 | 17.38  (16.63; 18.13) |
